# Epidemiology and Clinical Characteristics in Individuals with Confirmed SARS-CoV-2 Infection During the Early COVID-19 Pandemic in Saudi Arabia

**DOI:** 10.1101/2021.07.13.21260428

**Authors:** Fatimah S. Alhamlan, Reem S. Almaghrabi, Edward B. Devol, Anwar B. Alotaibi, Saleh M. Alageel, Dalia A. Obeid, Basem M. Alraddadi, Sahar I. Althawadi, Maysoon S. Mutabagani, Ahmed A. Al-Qahtani

## Abstract

Severe acute respiratory syndrome coronavirus 2 (SARS-CoV-2) is the causative agent of the catastrophic coronavirus disease 2019 (COVID-19) global pandemic. This study aimed to provide epidemiologic and clinical characteristics of patients with confirmed COVID-19 in Saudi Arabia and to determine whether characteristic profiles differ between patients who are symptomatic vs. asymptomatic for the disease. The first 492 consecutive patients diagnosed with SARS-CoV-2 infection at King Faisal Specialist Hospital and Research Centre in Saudi Arabia between March and September 2020 were included in this study. An electronic case report form developed using REDCap was used to collect data for each patient, including demographic characteristics, virus exposure (travel history, and human and animal contact), vaccination history, comorbidities, signs and symptoms, laboratory and radiographic reports, cardiac workup, medications, treatment regimens, and patient outcome. This patient cohort was 54% male, with 20.4% aged more than 60 years, 19.9% aged 31 to 40 years, and 17% aged 41 to 50 years. Most patients (79.2%) were symptomatic. Variables that significantly differed between symptomatic and asymptomatic patients were age, blood oxygen saturation percentage, hemoglobin level, lymphocyte count, neutrophil to lymphocyte (NTL) ratio, alanine aminotransferase (ALT) level, and aspartate aminotransferase (AST) level. Asymptomatic patients were mostly younger, with lower body mass index and ALT and AST levels but higher lymphocyte counts, NTL ratio, and CD4, CD8, natural killer cell, IgG, and IgM levels. The median incubation period reported for this cohort was 16 day, with upper and lower 95% quartiles of 27 and 10 days, respectively. Factors associated with increased risk of mortality were age (older than 42 years) and comorbidities, including specifically diabetes mellitus and hypertension. Patients who were not given an antiviral regimen were associated with better prognosis than patients who received an antiviral regimen (HR, 0.07; 95% CI, 0.011-0.25). Similar to countries worldwide, Saudi Arabia has explored treatment options to save the lives of patients during the COVID-19 pandemic. Our analyses will inform clinicians as well as policy makers to adopt the best strategies for SARS-CoV-2 infection management and treatment options.

## 1. INTRODUCTION

Soon after the novel corona virus, severe acute respiratory syndrome coronavirus 2 (SARS-CoV-2), emerged in Wuhan, China, in December 2019, its transmission from human to human established a foothold worldwide [1, 2]. In Saudi Arabia, the first case of a SARS-CoV-2– infected person was reported on March 2, 2020, a Saudi national who had returned to the country from Iran [3]. As of July 11, 2021, there have been over 500,000 confirmed cases, 481,241 recovered cases, and 7963 deaths in Saudi Arabia [4]. The number of confirmed positive cases had been escalating until now, July 2021, despite the country’s strict control measures.

Viral epidemiology and clinical manifestation play critical roles in the battle against SARS-CoV-2. Several studies have reported that patients infected with SARS-CoV-2 have primarily mild symptoms and good prognosis. Overall, a small fraction of infected patients have been reported to show severe pneumonia, pulmonary complications, acute respiratory distress syndrome, or multiple organ failure or to have died [2]. As of July 11, 2021, worldwide statistics showed a total of 185 million cases and 4 million deaths [5].

When the world was struck by this pandemic, hospitals leaders around the globe were unprepared for the scope of the infection. Indeed, there is still much to learn to manage the consequences of infections by this continually evolving virus. A lack of effective antiviral medication led to numerous clinical trials of different regimens using available and repurposed drugs to reach the best management of the disease caused by the virus, coronavirus 2019 (COVID-19). At King Faisal Specialist Hospital and Research Centre (KFSHRC), different treatment options following international and local guidelines and recommendations were offered to patients, including azithromycin, hydroxychloroquine (HCQ), azithromycin plus HCQ, interferon, lopinavir/ritonavir, lopinavir/ritonavir plus ribavirin, and azithromycin plus HCQ plus lopinavir/ritonavir [6-11].

The present study aimed to identify the epidemiologic characteristics, clinical features, laboratory test results, treatment regimens, and clinical outcomes for patients admitted to KFSHRC, a tertiary and referral hospital in Saudi Arabia, between March and September 2020, and to determine whether patient profiles differed significantly between patients who were symptomatic and those who were asymptomatic.

## 2. MATERIALS AND METHODS

### 2.1 Ethical Considerations

This study was conducted according to the World Medical Association Declaration of Helsinki and conforms to the ethics recommendations of the Committee on Publication Ethics and the International Committee of Medical Journal Editors. The study protocol was approved by the Research Advisory Council (Ethics Committee) at KFSHRC (RAC #220 1047), which also waived the requirement for obtaining informed patient consent because the risk to individuals whose data were included in the study was minimal and the study used exclusively retrospective deidentified administrative records.

### 2.2 Data Collection

Data regarding patients with confirmed COVID-19 (positive SARS-CoV-2 polymerase chain reaction results) were obtained from KFSHRC electronic medical records from March to September 2020. Data collectors were trained to ensure the completeness and accuracy of the data collected. An electronic case report form was developed using REDCap (a secure and flexible web-based clinical research data capture platform) that had over 500 data items for each patient, including demographic characteristics, potential virus exposure (travel history and human and animal contact), vaccination history, comorbidities, signs and symptoms, laboratory and radiologic reports, cardiac workup, medication, treatment regimen, and patient outcome. An unambiguous identification code was used that enabled identification of all data reported for each patient. The risk to study participants was limited to the potential loss of confidentiality. Appropriate measures were taken to prevent loss of participant confidentiality, including storage of electronic case report forms in a secure manner and presentation of data without identification of individual patients.

### 2.3 Data and Statistical Analyses

All collected data were stored and analyzed using SAS, version 9.4, software and GraphPad, version 9.0 (Prisma). Inferential and descriptive statistics were conducted to assess the epidemiologic and clinical manifestations of SARS-CoV-2 infections in Saudi Arabia. We performed *t* tests and Mann-Whitney tests to assess continuous variables, and χ^2^ tests to assess categorical variables. Univariate and multivariate analyses were performed using Cox proportional hazards regression models to identify factors associated with death from COVID-19. The hazard ratio (HR) along with the 95% confidence interval (CI) are reported. All reported *P-*values were two-tailed and were considered to be statistically significant at <0.05. Logistic regression models were used to assess the prediction value of immunological parameters.

For estimating the time-varying reproduction number, we used the EpiEstim package, version 2.2.1, with R software, version 3.3 [12]. The model estimated the time-varying reproduction number from the daily number of cases of COVID-19 reported at KFSHRC and an uncertain generation time with a mean of 4.6 days and a standard deviation (SD) of 2.9 days based on other studies [13]. We used a gamma distribution prior for the reproduction number with a mean of 2.6 and SD of 2.0 based on early estimates for the basic reproduction number (R0) from the initial stages [14].

## 3. RESULTS

### 3.1 Summary of Demographic Characteristics and Clinical Data

Data from 492 patients were collected from the portal from March 10 to September 11, 2020. Table 1 provides a summary of patient demographic and clinical characteristics for the cohort. Of 492 patients, 54.0% were male. Most patients (20.4%) were older than 60 years, followed by ages ranging from 31 to 40 years (20.0%) and from 41 to 50 years (17.9%). The highest proportion of patients in this cohort received at KFSHRC were from Saudi Arabia (77.8%), followed by the Philippines (29.4%), and India (22.4%). Of those from Saudi Arabia, the majority were from Riyadh (79%), followed by Jeddah (15.1%). The clinical data indicated that the majority of patients symptomatic for COVID-19 presented with a temperature lower than 38 °C (54.8%) and without a dry cough (51.6%). Only 21.2% reported myalgia fatigue, and 23.6% had chest radiographs with indications of concern. Intensive care unit (ICU) admission was received by 21.8% of the cohort, and 12.1% received mechanical ventilation. We found that 42.0% of patients presented with comorbidities, including 20.6% with diabetes and 24.4% with hypertension. For patient outcomes on day 14 of hospitalization, 33.2% showed persistent disease, 16.1% recovered, 8.4% were discharged from the hospital, and 1.5% died.

**Table 1.** Demographic and clinical characteristics of 492 patients.

| Characteristic | No. of patients | Percentage of total patients | $\chi^2$ (P-value) |
| --- | --- | --- | --- |
| **Age** |  |  | 16.23 (0.01) |
| <20 | 66 | 13.4 |  |
| 21-30 | 61 | 12.4 |  |
| 31-40 | 98 | 19.9 |  |
| 41-50 | 88 | 17.9 |  |
| 51-60 | 78 | 15.9 |  |
| >60 | 100 | 20.4 |  |
| **Sex** |  |  | 3.1 (0.076) |
| Male | 262 | 54.0 |  |
| Female | 223 | 45.9 |  |
| Unknown | 6 | NA* |  |
| **Nationality** |  |  |  |
| Saudi | 372 | 77.8 | 148 (<0.0001) |
| Non-Saudi | 106 | 22.2 |  |
| Unknown | 13 | NA* |  |
| **Other nationalities** |  |  |  |
| Bangladesh | 6 | 7.1 |  |
| British | 2 | 2.4 |  |
| Canadian | 1 | 1.2 |  |
| Egyptian | 2 | 2.4 |  |
| Filipino | 25 | 29.4 |  |
| Indian | 19 | 22.4 |  |
| Jordanian | 7 | 8.2 |  |
| Lebanese | 1 | 1.2 |  |
| Nigerian | 1 | 1.2 |  |
| Pakistani | 9 | 10.6 |  |
| Sudanese | 7 | 8.2 |  |
| Swiss | 1 | 1.2 |  |
| Syrian | 1 | 1.2 |  |
| Yemeni | 1 | 1.2 |  |
| **Saudi city or region** |  |  |  |
| Riyadh | 377 | 79 | 3541 (0.0001) |
| Jeddah | 72 | 15.1 |  |
| Eastern Region | 10 | 2.1 |  |
| Ahsa | 2 | 0.4 |  |
| Albaha | 1 | 0.2 |  |
| Asir | 2 | 0.4 |  |
| Hail | 1 | 0.2 |  |
| Jazan | 1 | 0.2 |  |
| Madinah | 1 | 0.2 |  |
| Najran | 3 | 0.6 |  |
| Northern Borders | 3 | 0.6 |  |
| Tabouk | 2 | 0.4 |  |
| Unknown | 14 | NA* |  |
| **Smoking status** |  |  | 223 (<0.0001) |
| Smoker | 43 | 11.4 |  |
| Nonsmoker | 333 | 88.6 |  |
| Unreported | 115 | NA* |  |
| **Pregnant** |  |  | 177 (<0.001) |
| Yes | 11 | 5.02 |  |
| No | 208 | 94.9 |  |
| Unreported | 272 | NA* |  |
| **HCW** |  |  | 32 (<0.0001) |
| Yes | 110 | 23.2 |  |
| No | 169 | 35.6 |  |
| Unknown | 212 | NA* |  |
| **BMI (kg/m<sup>2</sup>)** |  |  | 168 (<0.0001) |
| <18.5 | 166 | 49.3 |  |
| 19-30 | 7 | 1.9 |  |
| 25-30 | 56 | 15.9 |  |
| >30 | 122 | 34.8 |  |
| Unknown | 140 | NA* |  |
| **Body temperature >38 °C** |  |  | 261 (<0.001) |
| Yes | 154 | 32.9 |  |
| No | 257 | 54.8 |  |
| Not assessed | 58 | 12.3 |  |
| **Dry Cough** |  |  |  |
| Yes | 152 | 32.1 | 126 (<0.001) |
| No | 244 | 51.6 |  |
| Not assessed | 77 | 16.3 |  |
| **Runny Nose** |  |  |  |
| Yes | 72 | 15.4 | 174 (<0.001) |
| No | 289 | 61.7 |  |
| Not assessed | 107 | 22.9 |  |
| **Myalgia fatigue** |  |  |  |
| Yes | 113 | 21.2 | 82 (<0.001) |
| No | 248 | 53.1 |  |
| Not assessed | 106 | 22.7 |  |
| **Chest Radiograph** |  |  | 56 (<0.001) |
| Normal | 231 | 49.6 |  |
| Abnormal | 110 | 23.6 |  |
| Not assessed | 125 | 26.8 |  |
| **Intensive care unit** |  |  |  |
| Yes | 100 | 21.8 | 222.2 (<0.001) |
| No | 301 | 65.7 |  |
| Not assessed | 33 | NA* |  |
| **Mechanical ventilation** |  |  |  |
| Yes | 55 | 12.1 | 327.5 (<0.001) |
| No | 333 | 73.4 |  |
| Unknow | 103 | NA* |  |
| **Comorbidities** |  |  |  |
| Yes | 206 | 41.9 | 12 (<0.0001) |
| No | 285 | 58.0 |  |
| **Diabetes mellitus** |  |  |  |
| Yes | 101 | 20.5 | 170 (<0.0001) |
| No | 390 | 79.4 |  |
| **Hypertension** |  |  | 128 (<0.0001) |
| Yes | 120 | 24.4 |  |
| No | 371 | 75.6 |  |
| **Outcome on day 14** |  |  |  |
| Recovered | 65 | 16.1 | 220 (<0.001) |
| Death | 6 | 1.5 |  |
| Discharge | 34 | 8.4 |  |
| Persistent disease | 134 | 33.2 |  |
| Not documented | 252 | NA * |  |
| **Travel to countries outside Saudi Arabia** |  |  | 218.9 (<0.0001) |
| Yes | 31 | 6.5 |  |
| No | 293 | 62.1 |  |
| Not reported | 133 | NA* |  |

Abbreviations: BMI, body mass index; HCW, health care worker.
\*Not included in the statistical test.

### 3.2 Demographic and Clinical Characteristics of Symptomatic vs. Asymptomatic Patients

The second aim of our study was to determine the demographic and clinical characteristics of symptomatic vs. asymptomatic persons. Of 429 individuals in the cohort, 79.2% reported being symptomatic and 20.7% being asymptomatic. A summary of the analysis assessing continuous variables by symptomatic status is given in Table 2. Characteristics that were significantly different between symptomatic and asymptomatic patients were age, blood oxygen saturation percentage, hemoglobin level, lymphocyte level, the neutrophil to lymphocyte (NTL) ratio, alanine aminotransferase (ALT) level, and aspartate aminotransferase (AST) level. Asymptomatic patients were primarily younger, with lower body mass index and ALT and AST levels, but higher NTL ratios and higher lymphocyte, CD4, CD8, natural killer cell, IgG, and IgM levels.

**Table 2.**
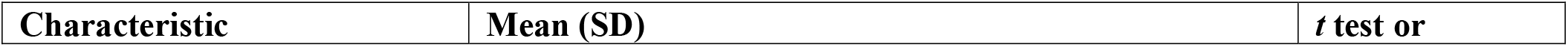
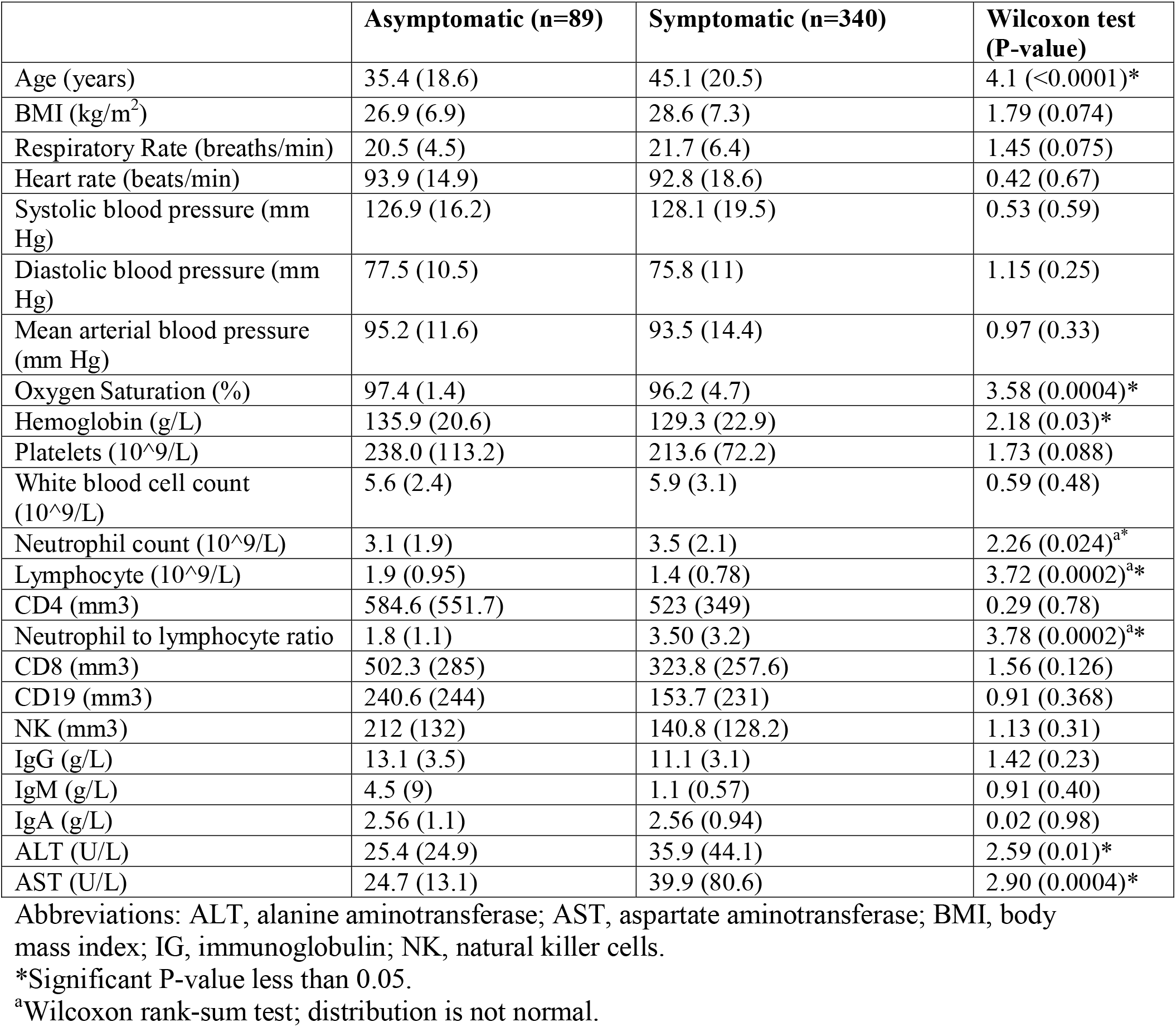
Continuous demographic and clinical characteristics assessed by symptomatic vs. asymptomatic status.

| Characteristic | Mean (SD) | t test or |
| --- | --- | --- |

|  | Asymptomatic (n=89) | Symptomatic (n=340) | Wilcoxon test (P-value) |
| --- | --- | --- | --- |
| Age (years) | 35.4 (18.6) | 45.1 (20.5) | 4.1 (<0.0001)* |
| BMI (kg/m<sup>2</sup>) | 26.9 (6.9) | 28.6 (7.3) | 1.79 (0.074) |
| Respiratory Rate (breaths/min) | 20.5 (4.5) | 21.7 (6.4) | 1.45 (0.075) |
| Heart rate (beats/min) | 93.9 (14.9) | 92.8 (18.6) | 0.42 (0.67) |
| Systolic blood pressure (mm Hg) | 126.9 (16.2) | 128.1 (19.5) | 0.53 (0.59) |
| Diastolic blood pressure (mm Hg) | 77.5 (10.5) | 75.8 (11) | 1.15 (0.25) |
| Mean arterial blood pressure (mm Hg) | 95.2 (11.6) | 93.5 (14.4) | 0.97 (0.33) |
| Oxygen Saturation (%) | 97.4 (1.4) | 96.2 (4.7) | 3.58 (0.0004)* |
| Hemoglobin (g/L) | 135.9 (20.6) | 129.3 (22.9) | 2.18 (0.03)* |
| Platelets (10<sup>9</sup>/L) | 238.0 (113.2) | 213.6 (72.2) | 1.73 (0.088) |
| White blood cell count (10<sup>9</sup>/L) | 5.6 (2.4) | 5.9 (3.1) | 0.59 (0.48) |
| Neutrophil count (10<sup>9</sup>/L) | 3.1 (1.9) | 3.5 (2.1) | 2.26 (0.024)<sup>a*</sup> |
| Lymphocyte (10<sup>9</sup>/L) | 1.9 (0.95) | 1.4 (0.78) | 3.72 (0.0002)<sup>a*</sup> |
| CD4 (mm3) | 584.6 (551.7) | 523 (349) | 0.29 (0.78) |
| Neutrophil to lymphocyte ratio | 1.8 (1.1) | 3.50 (3.2) | 3.78 (0.0002)<sup>a*</sup> |
| CD8 (mm3) | 502.3 (285) | 323.8 (257.6) | 1.56 (0.126) |
| CD19 (mm3) | 240.6 (244) | 153.7 (231) | 0.91 (0.368) |
| NK (mm3) | 212 (132) | 140.8 (128.2) | 1.13 (0.31) |
| IgG (g/L) | 13.1 (3.5) | 11.1 (3.1) | 1.42 (0.23) |
| IgM (g/L) | 4.5 (9) | 1.1 (0.57) | 0.91 (0.40) |
| IgA (g/L) | 2.56 (1.1) | 2.56 (0.94) | 0.02 (0.98) |
| ALT (U/L) | 25.4 (24.9) | 35.9 (44.1) | 2.59 (0.01)* |
| AST (U/L) | 24.7 (13.1) | 39.9 (80.6) | 2.90 (0.0004)* |

Abbreviations: ALT, alanine aminotransferase; AST, aspartate aminotransferase; BMI, body mass index; IG, immunoglobulin; NK, natural killer cells.
\*Significant P-value less than 0.05.
<sup>a</sup>Wilcoxon rank-sum test; distribution is not normal.

Table 3 provides a summary of categorical data by symptom status. Many demographic and clinical characteristics were significantly different between the two groups, including, age, sex, smoking status, influenza vaccine receipt, comorbidities, diabetes, hypertension, chest radiographic findings, pneumonia treatment, and outcome on days 7 and 14. The highest proportion of asymptomatic patients were from age group 31–40 years and were men. Most asymptomatic patients were nonsmokers, had received an influenza vaccine, and had no history of comorbidities, including diabetes or hypertension.

**Table 3.**
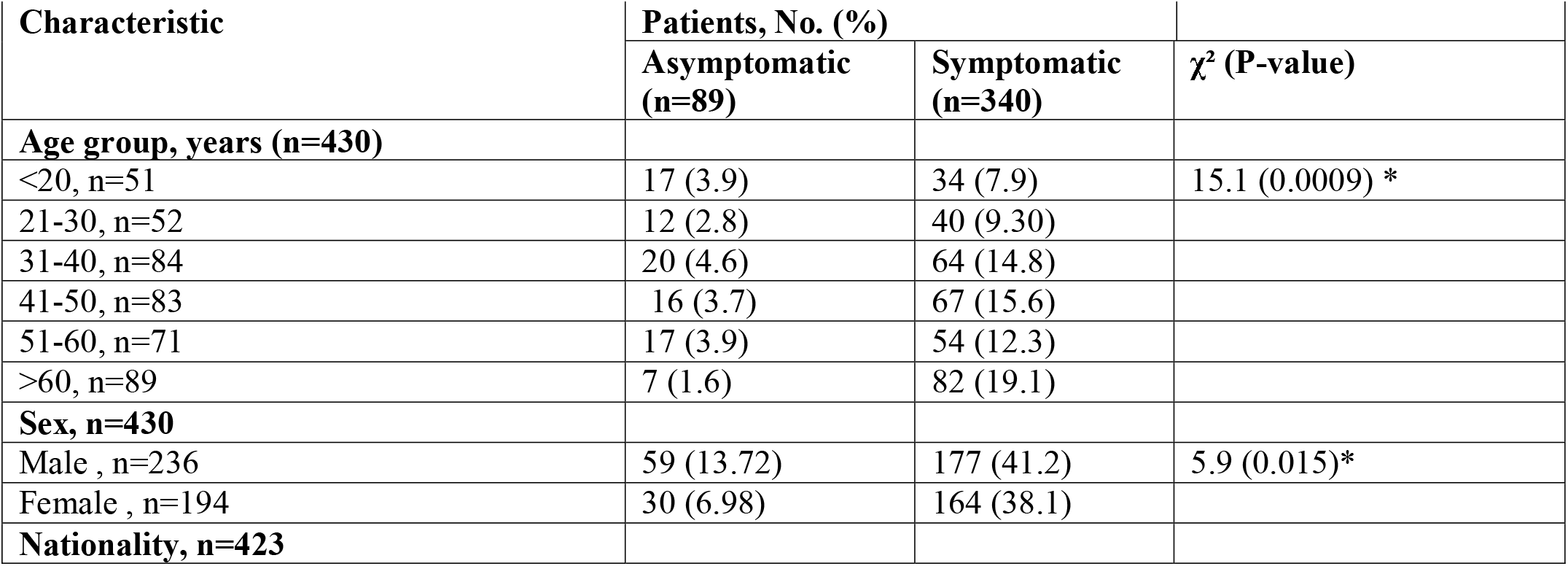
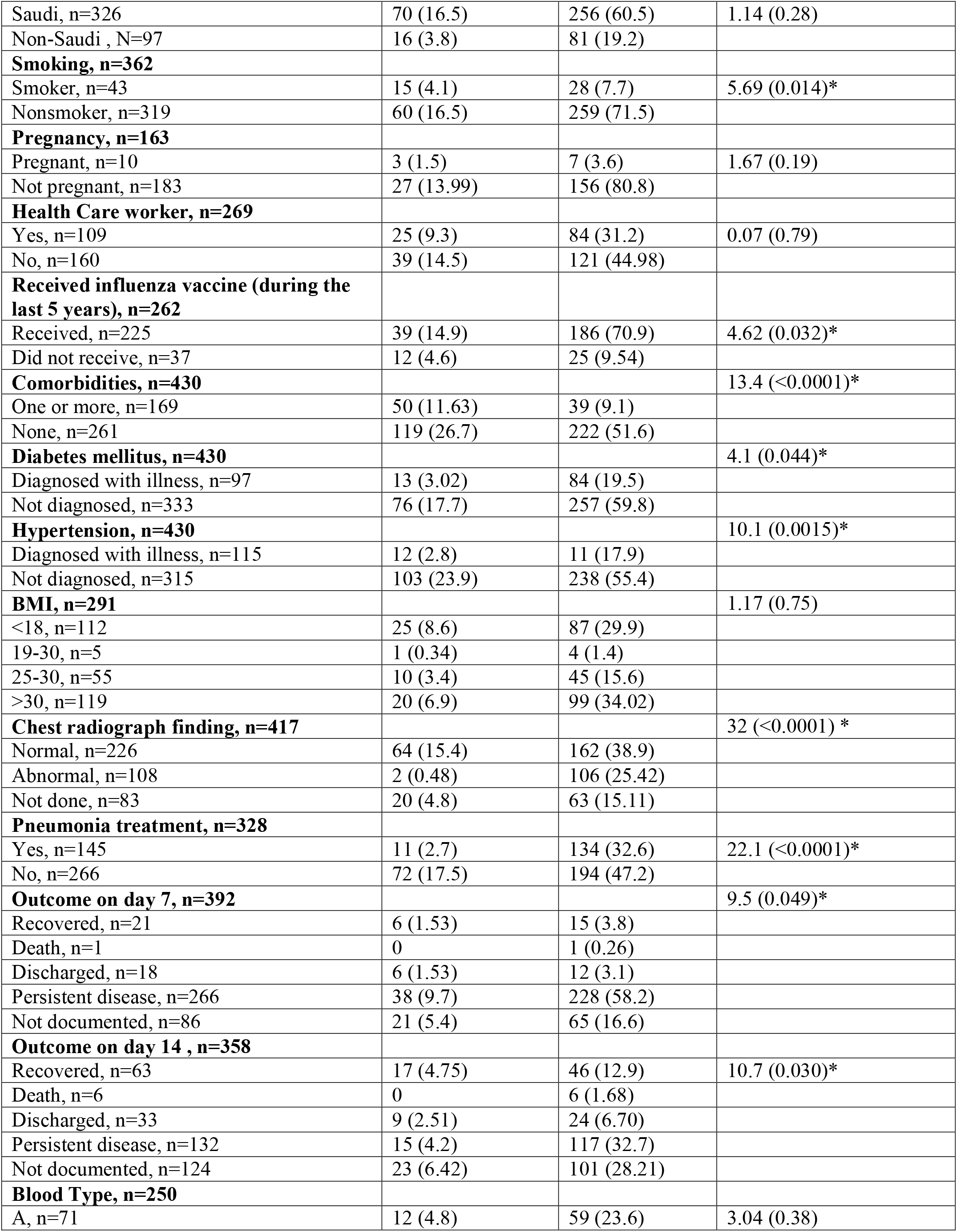
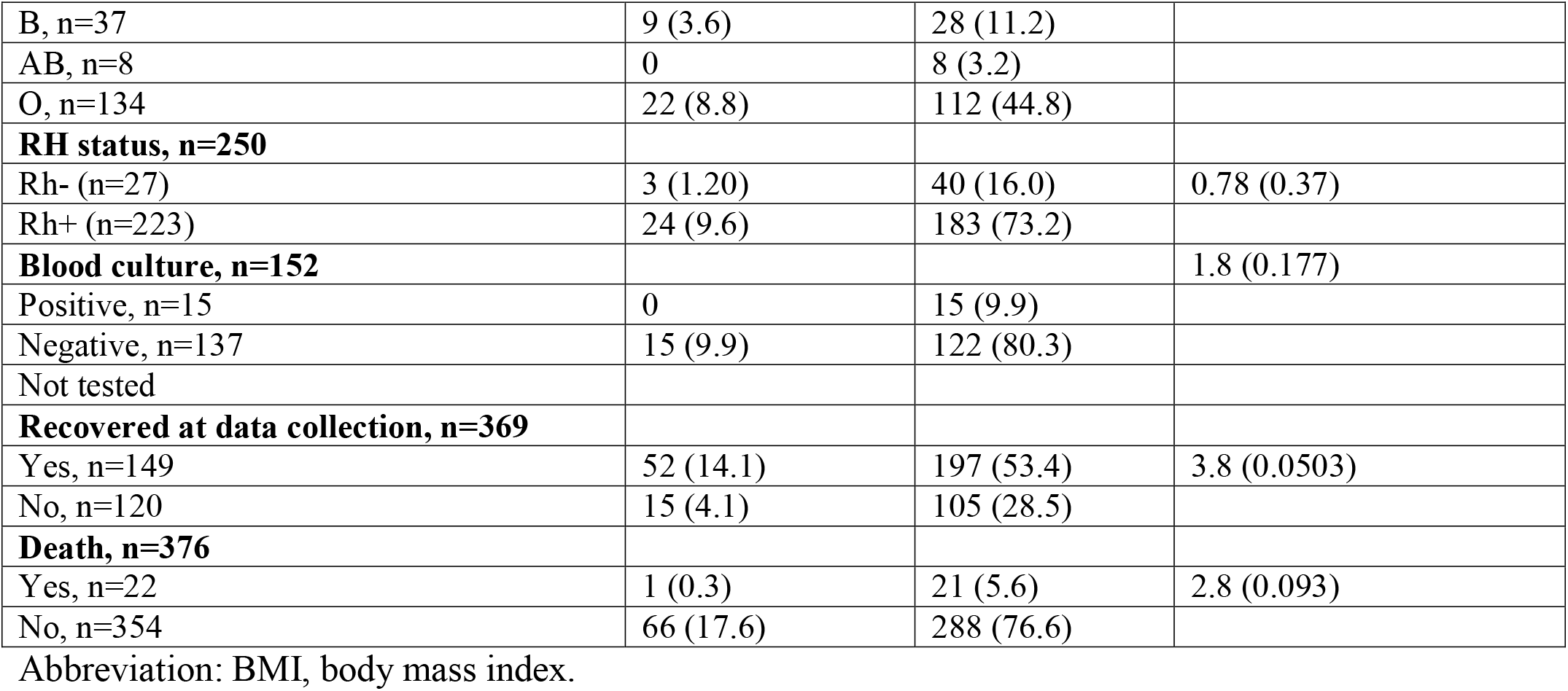
Categorical demographic and clinical characteristics assessed by symptomatic vs. asymptomatic status

### 3.3 Variation in Disease Incubation Times

The median incubation period reported in our population was 16 days (SD, 17 days), the upper and lower 95% quartiles were 27 and 10 days, respectively. The incubation period differed significantly only with age: patients 42 years or younger reported lower incubation times than those older than 42 years. The incubation period did not differ significantly between any other examined demographic or clinical variable, including the number of treatment regimens (Table 4).

**Table 4.** Incubation time differences by demographic or clinical characteristic or treatment regimen

| Characteristic | Incubation time, mean (SD), days | T test (P-value ) |
| --- | --- | --- |
| **Age** |  |  |
| ≤42 | 19.9 (20.2) | 2.25 (<0.0001)* |
| >42 | 24.3 (20.2) |  |
| **Sex** |  |  |
| Male | 21.1 (17.5) | 0.44 (0.66) |
| Female | 22.2 (17.2) |  |
| **Nationality** |  | 1.31 (0.097) |
| Saudi | 20.6 (16.8) |  |
| Non-Saudi | 25.5 (19.2) |  |
| **Smoking status** |  |  |
| Smoker | 18.5 (10.8) | 0.45 (0.657) |
| Nonsmoker | 19.8 (16.5) |  |
| **Symptom Status** |  |  |
| Symptomatic | 20.9 (16.1) |  |
| Asymptomatic | 20.75 (20.5) | 0.07 (0.94) |
| **Treatment regimen** |  |  |
| One or none | 22.85 (18.4) | 1.06 (0.28) |
| More than two | 20.2 (15.9) |  |
| **Comorbidities** |  |  |
| One or more | 23.4 (19) | 1.1 (0.27) |
| None | 20.5 (16.15) |  |
| **Diabetes mellitus** |  |  |
| Diagnosed with illness | 23.8 (15.6) | 0.80 (0.43) |
| Not diagnosed | 21.1 (17.1) |  |
| **Hypertension** |  | 1.1 (0.29) |
| Diagnosed with illness | 23.8 (25.6) |  |
| Not diagnosed | 20.9 (23.7) |  |

\*Significant P-value less than 0.05.

### 3.4 Demographical and Clinical Characteristics Associated with Death Due to COVID-19

The epidemiologic and clinical factors associated with death among patents with COVID-19 are summarized in Table 5. The only demographic characteristic that was statistically significantly associated with death for patients with COVID-19 was being older than 42 years (HR, 10.32; 95% CI, 2.4-44.3) although the risk of death appeared higher for men than women, Saudi nationals vs. non-Saudi persons, and individuals with vs. without obesity. Assessment of clinical data indicated that being symptomatic or asymptomatic was not significantly associated with higher risk of death, whereas having comorbidities in general was significantly associated with higher risk of death (HR, 3.42; 95% CI, 1.24-12.1) as was specifically having diabetes mellitus (HR, 3.9; 95% CI, 1.68-9.4) or hypertension (HR, 5.135; 95% CI, 2.2-12.5). None of the common complications of COVID-19 (e.g., fever, myalgia fatigue, sore throat, and vomiting) were significantly associated with death, although among these complications, higher risk of poor prognosis was observed for patients with chest radiograph results indicating abnormalities, followed by patients with dry cough and nausea. Regarding treatment options, patients who were not given an antiviral regimen had better prognoses (HR, 0.07; 95% CI, 0.011-0.25) than patients who received an antiviral regimen, followed by patients receiving interferon regimens. Significant and high risk of death was associated with receipt of a combination therapy of azithromycin, hydroxychloroquine, and lopinavir/ritonavir (HR, 149.6; 95% CI, 5.8-3808.4), whereas lower risk of death was associated with receipt of a combination of two or more therapies (HR, 2.7; 95% CI, 1.1-6.6). For receipt of a single treatment, among those examined, lopinavir/ritonavir (HR, 19.28; 95% CI, 0.98-130.4) and azithromycin (HR, 3.5; 95% CI, 0.819-10.4) were associated with the worst prognosis. Patients receiving pneumonia treatment were associated with poor prognosis (HR, 9.1; 95% CI, 3.28-32.3), and patients receiving mechanical ventilation had the second highest HR (HR, 90; 95% CI, 18.4-1624.2).

**Table 5.** Association of demographic and clinical characteristics and treatment regimen with death in patients with confirmed COVID-19

| **Variable** | **Cox regression univariate model HR** | **95% Confidence interval** | **P-value** |
| --- | --- | --- | --- |
| **Sex** |  |  |  |
| Female | 1(Ref) |  |  |
| Male | 1.56 | 0.67-3.66 | 0.30 |
| **Age** |  |  |  |
| ≤42 | 1(Ref) |  |  |
| >42 | 10.32 | 2.4-44.3 | 0.0017* |
| **Nationality** |  |  |  |
| Saudi | 1.93 | 0.557-12.2 | 0.37 |
| Non-Saudi | 1(Ref) |  |  |
| **BMI (kg/m<sup>2</sup>)** |  |  |  |
| ≤30 | 1(Ref) |  |  |
| >30 | 2.28 | 0.91-5.44 | 0.065 |
| **Symptom status** |  |  |  |
| Asymptomatic | 1(Ref) |  |  |
| Symptomatic | 4.68 | 0.942-85 | 0.137 |
| **Comorbidities** |  |  |  |
| Reported | 3.42 | 1.24-12.1 | 0.0293* |
| Not reported | 1(Ref) |  |  |
| **Diabetes mellitus** |  |  |  |
| Reported | 3.9 | 1.68-9.4 | 0.0015* |
| Not reported | 1(Ref) |  |  |
| **Hypertension** |  |  |  |
| Reported | 5.135 | 2.2-12.5 | 0.0002* |
| Not reported | 1(Ref) |  |  |
| **Comorbidities related to autoimmune disorders<sup>(a)</sup>** |  |  | 0.0093* |
| Reported | 3.1 | 1.3-7.4 |  |
| Not reported | 1(Ref) |  |  |
| **Complications (Ref = no complication)** |  |  |  |
| Abdominal pain | 1.67 | 0.35-4.95 | 0.48 |
| Chest radiograph (abnormal findings) | 5.4 | 1.1-97.7 | 0.102 |
| Dry cough | 2.57 | 0.97-6.92 | 0.0532 |
| Fever (>38 °C) | 1.79 | 0.72-4.63 | 0.21 |
| Myalgia fatigue | 2.26 | 0.69-7.11 | 0.1662 |
| Nausea | 2.48 | 0.67-7.62 | 0.132 |
| Productive cough | 1.77 | 0.56-4.87 | 0.29 |
| Runny nose | 0.94 | 0.15-3.45 | 0.93 |
| Sore throat | 1.18 | 0.37-3.34 | 0.76 |
| Vomiting | 2.1 | 0.45-7.1 | 0.28 |
| **Treatment Option** |  |  |  |
| Not given a specific drug regimen | 0.07 | 0.011-0.25 | 0.0005* |
| Given a specific drug regimen | 1(Ref) |  |  |
| Azithromycin | 3.5 | 0.819-10.4 | 0.045* |
| Not given | 1(Ref) |  |  |
| HCQ | 1.52 | 0.24-5.23 | 0.575 |
| Not given | 1(Ref) |  |  |
| Interferon | 0 | 0-154 | 0.993 |
| Not given | 1(Ref) |  |  |
| lopinavir/ritonavir | 19.28 | 0.98-130.4 | 0.0008* |
| Not given | 1(Ref) |  |  |
| Azithromycin + HCQ | 2.21 | 0.93-5.32 | 0.071 |
| Not given | 1(Ref) |  |  |
| lopinavir/ritonavir + Ribavirin | 0 | 0-50.7 | 0.997 |
| Not given | 1(Ref) |  |  |
| Azithromycin + HCQ + lopinavir/ritonavir | 149.6 | 5.8-3808.4 | 0.0004* |
| Not given | 1(Ref) |  |  |
| Pneumonia Treatment | 9.1 | 3.28-32.3 | 0.0001* |
| Not given | 1(Ref) |  |  |
| Combination Therapy (2 or more) | 2.7 | 1.1-6.6 | 0.026* |
| Not given | 1(Ref) |  |  |
| Received mechanical ventilation | 90 | 18.4-1624.2 | <0.0001* |
| Not received | 1(Ref) |  |  |

Abbreviation: BMI, body mass index; HCQ, Hydroxychloroquine
<sup>a</sup>Comorbidity linked to immunity dysfunction including diabetes, rheumatoid, lupus IBS, and APS
\*Significant P-value less than 0.05.

### 3.5 Association of Immunological Factors with Patient Outcomes, Symptoms, and Treatment Regimen

Using multivariate Cox regression, we evaluated white blood cell (WBC), absolute lymphocyte, and neutrophil counts and the NTL ratio as factors potentially associated with patient outcomes and with type of treatment regimen. The values of these factors were obtained from patients after they received a diagnosis of COVID-19. If a univariate model indicated that a factor was significantly associated with patient outcome, we pursued additional testing for treatment type. The results of a univariate Cox regression assessing WBC count indicated a significant association with patient outcome (χ^2^=11.9, P=0.0005). The multivariate model testing treatment types showed significance for all treatment types (P<0.05). For patients receiving lopinavir/ritonavir treatment alone, the HR was significant (HR=35.3; 95% CI, 1.7-312.4), whereas the HR for WBC count was 1.2 (95% CI, 1.1-1.3). The other significant HR was for receipt of combination therapy with lopinavir/ritonavir, HCQ, and azithromycin (HR=154.5; 95% CI, 5.9-4013). For Patients receiving treatment with HCQ or azithromycin alone or with the combination of HCQ and azithromycin or with interferon or for pneumonia, HRs were not statistically significant (P>0.05).

A univariate cox regression model assessing the association of lymphocyte count with patient outcome showed no significance (χ^2^=0.39, P=0.53). In addition, none of the multivariate models were significant (P>0.05). Similarly, none of the models indicated an association with the NTL ratio.

By contrast, for the neutrophil count, a univariate cox regression model assessing the association with patient outcome was statistically significant (χ^2^=6.7, P=0.0092). The multivariate models assessing an association with treatment types were significant for all treatment types (P<0.05). However, the only significant treatment factors were lopinavir/ritonavir and the combination of HCQ, azithromycin, and lopinavir/ritonavir. For patients receiving lopinavir/ritonavir treatment alone, the HR was significant (HR=23.5; 95% CI, 1.1-196), whereas the neutrophil HR was 1.3 (95% CI, 1.1-1.5). For patients receiving the combination therapy, the HR was significant (HR, 92.8; 95% CI, 3.6-2348), whereas the neutrophil HR was 1.3 (95% CI, 1.1-1.5). For patients receiving HCQ treatment, interferon treatment, any two-drug combination therapy, or pneumonia treatment, the HRs were not significant (P>0.05).

We also evaluated models assessing associations of levels of immunological factors with patient symptom status. Neither the WBC levels nor the neutrophil count were associated with patient symptoms status. An immunological parameter that was significantly associated with patient symptom status was the NTL ratio (χ^2^=17.3, P<0.0001); higher NTL ratios were correlated with symptomatic disease (OR=0.6, 95% CI: 0.4-0.8). There was also a significant association between patient symptom status and lymphocyte levels alone (F=17.9, P <0.0001; R^2^=0.058); higher WBC count was correlated with asymptomatic disease.

We evaluated the use of a mechanical ventilator as a proxy for the severity of disease and found that it was significantly associated with the NTL ratio (χ^2^=14.9, P <0.0001, OR=1.2, 95% CI:1.1-1.3), with higher NTL ratios correlating with severe disease. Similarly, higher WBC counts (χ^2^=42.9, P<0.0001, OR=1.4, 95% CI:1.2-1.6) and higher neutrophil counts (χ^2^=28.5, P<0.0001, OR=1.5, 95% CI:1.3-1.8). were correlated with greater disease severity. Although higher lymphocyte levels appeared to be associated with less severe disease, the association was not significant (χ^2^=2.3, P= 0.09).

For patient outcomes, the model assessing the association of the WBC count (χ^2^=13.3, P=0.003, OR=1.2, 95% CI:1.2-1.4) and that assessing the level of neutrophils (χ^2^=11.0, P=0.0009, OR=1.4, 95% CI=1.4-1.7). were significant, with higher levels correlated with patient death. However, neither the NTL ratio nor lymphocyte level was associated with patient death. Figure 2 shows the distribution of the immunological factors by disease severity and tested by the Mann-Whitney test. Three multilevel logistic models were assessed with all immunological parameters, and they were all significant predictor for patient’s outcome, or severity or symptoms together (P<0.005).

**Figure 1.**
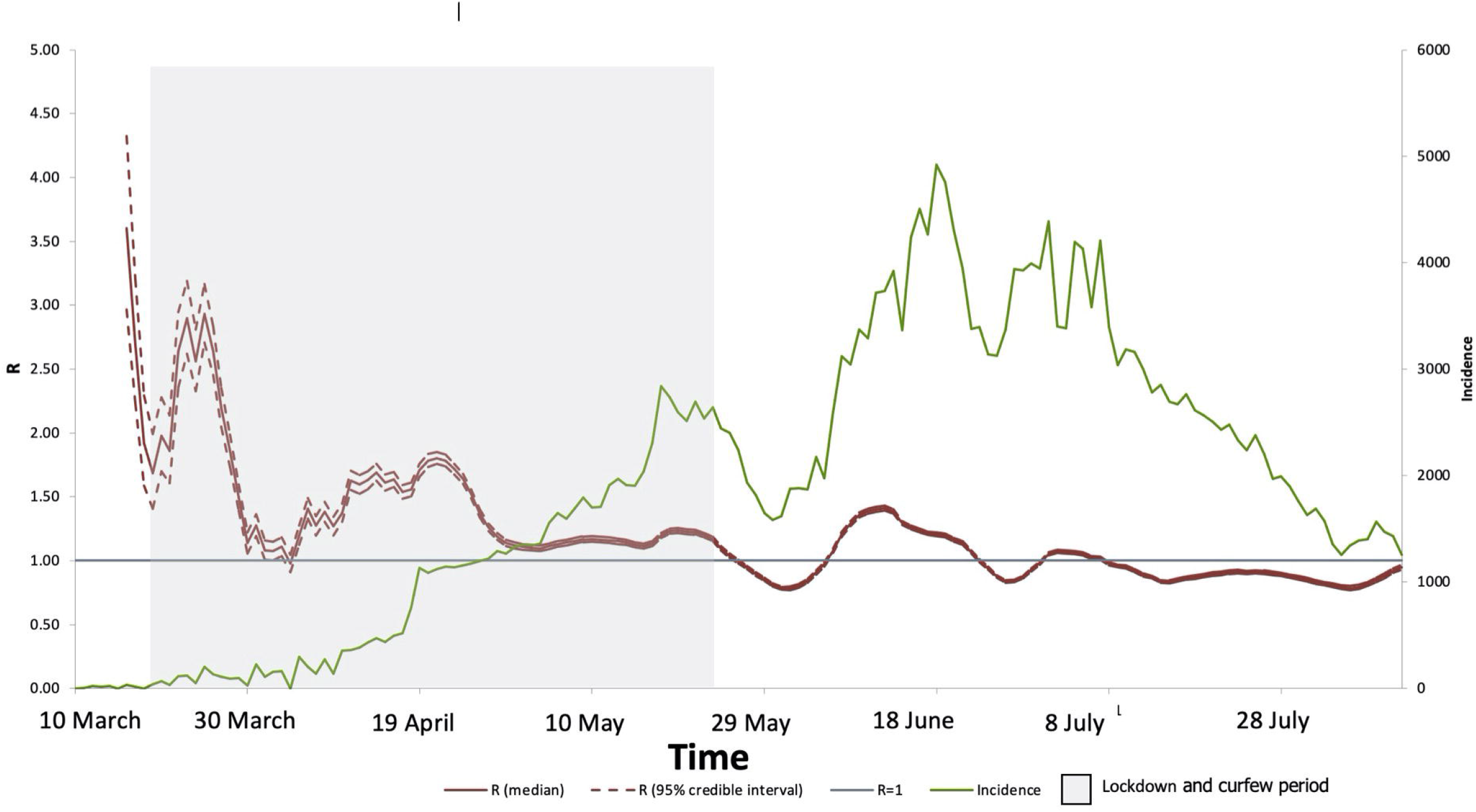
Age group distribution of COVID-19 cases by symptom status (asymptomatic vs. symptomatic) shown as percentages. Most patients in this cohort with confirmed COVID-19 were older than 60 years (19%); 21% of the cohort was asymptomatic, whereas 79% was symptomatic. The highest percentage of asymptomatic patients (5%) was observed in the age group of 30 to 39 years.

**Figure 2.**
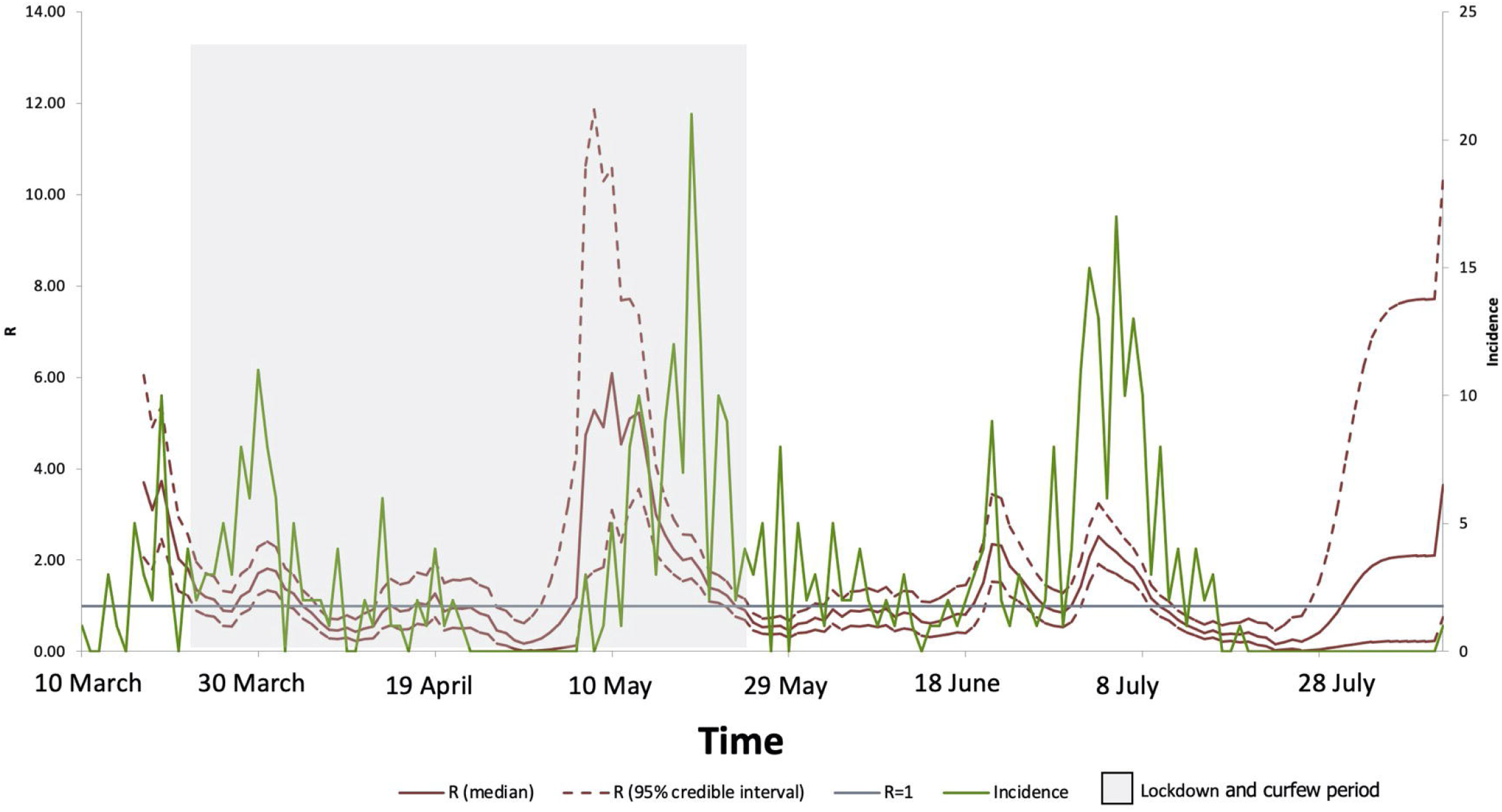
Box plots of patient blood test results after diagnosis by illness severity as assessed by the need for mechanical ventilation (MV). A) White blood cell count by illness severity, with higher counts observed for more severe illness. B) Neutrophil absolute count by illness severity, with higher counts observed for more severe illness. C) Lymphocyte absolute count by illness severity, with higher counts observed for milder illness. D) Neutrophil to lymphocyte (NTL) ratio by illness severity, with higher ratios observed for more severe illness.

A multivariate model using multilevel regression was used to assess the association of immunological factors with the incubation period. The lymphocyte count with age was the only factor significantly associated with the incubation period (F=4.8, P=0.009). Neutrophil and WBC counts and the NTL ratio were not associated.

### 3.6 Reproduction Number Estimations and Predicting COVID-19 in Riyadh

We estimated the instantons temporal effective reproduction number (R_t_) using the EpiEstim package and modeled it using a gamma distribution. For 155 days, confirmed COVID-19 cases were detected at KFSHRC. Figure 3 shows the summary of the incidence and R values over time. The average value for R from March 11 to August 10, 2020, was 1.21. In March, R averaged 1.94, decreasing in April to 0.81, but increasing in May to 2.24, before decreasing in June to 1.07 and in July to 0.92. The highest value of R was found in May, which is when the government implemented strict lockdown protocols to substantially decrease the number of confirmed COVID-19 cases in June and July, accompanied by a significant decrease in the value of R. In August, a significant decrease in cases was observed followed by an increased risk of R followed its mean values reported. We also estimated the effective reproduction number over time in Saudi Arabia overall. The pattern across the entire country was similar to that estimated for KFSHRC (Figure 4). However, R peaked at a national level in March then decreased in May after the lockdown and kept decreasing in June and July. The average value of R in Saudi Arabia between March and July was 1.23.

**Figure 3.**
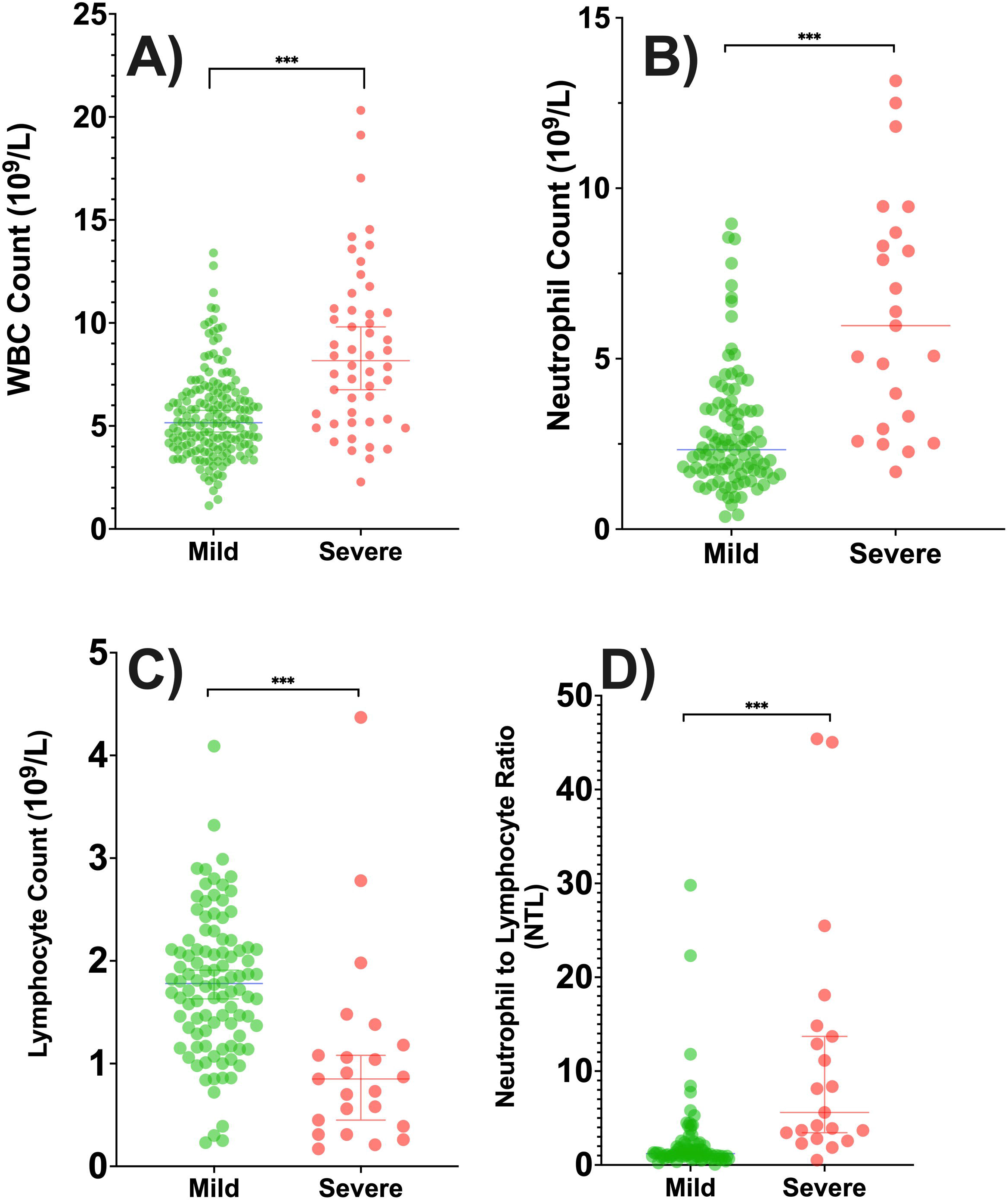
Instantons reproduction number (R_t_) estimated using the EpiEstim package and modeled using a gamma distribution. Cases were detected for 155 days at KFSHRC. The average R value from March 10 to August 11, 2020, was 1.21. The red line shows the average median of R; dashed red line, confidence intervals; green line, incidence of cases reported; gray area, the lockdown period; and blue line, R=1.

**Figure 4.**
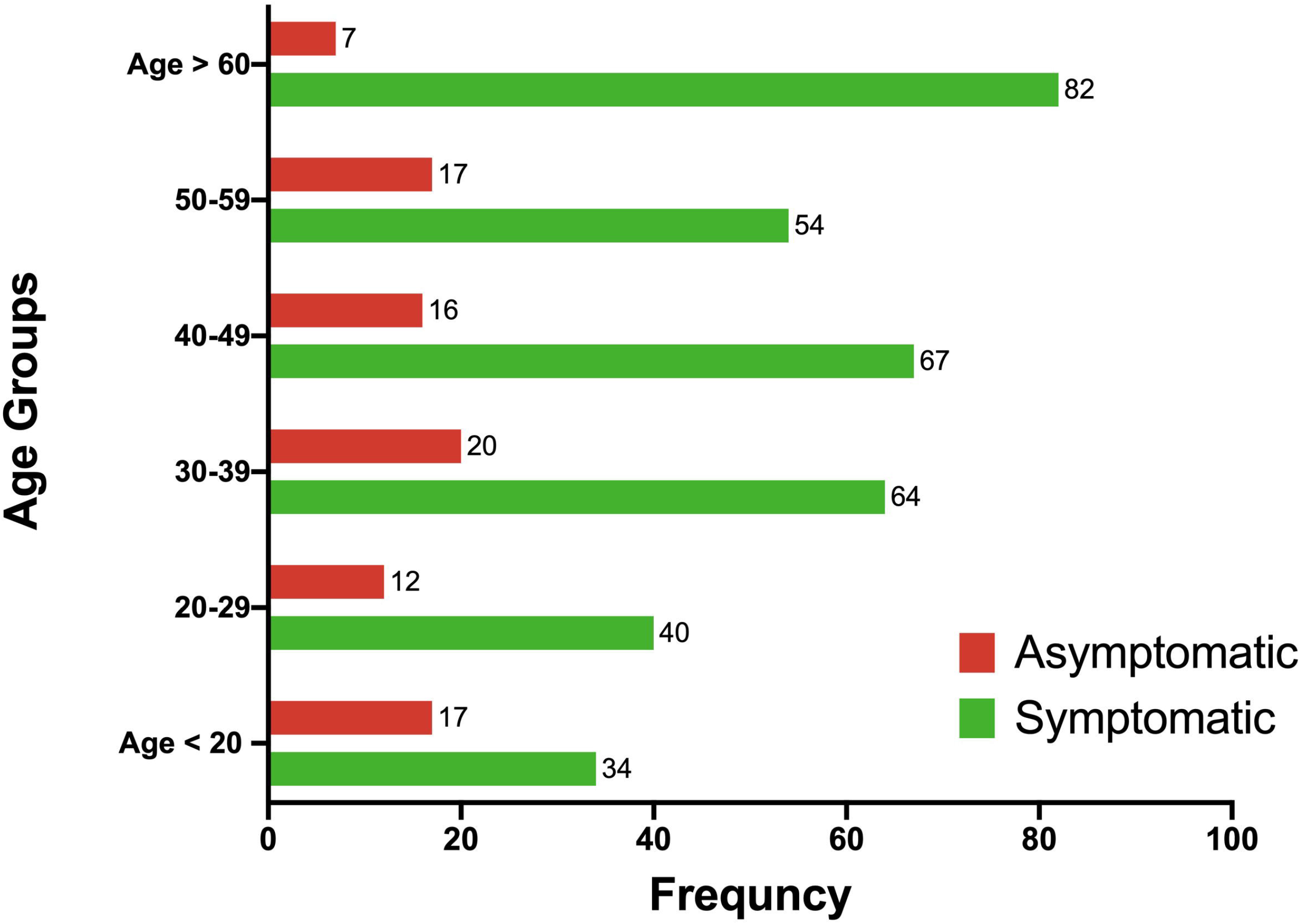
The estimated instantons reproduction number (R_t_), using EpiEstim package and model using gamma distribution. Cases were detected for 155 days in Saudi Arabia. The average R-value from March 10 to August 11, 2020, was 1.23. The red line shows the average median of R, the dashed red line shows the confidence intervals, while the green line shows the incidence of cases reported, the grey area represents the time of lockdown, while the blue line represents the R=1.

## 4. DISCUSSION

KFSHRC is a tertiary care hospital in Riyadh, Saudi Arabia, that typically provides specialized care for cancer, organ transplantation, and patients who are immunocompromised in addition to offering other medical specialties. However, to increase treatment capacity for Saudi Arabia during the pandemic, KFSHRC allowed access to COVID-19 patients when it is needed. The first case was admitted in March 2020. By investigating the demographic and clinical characteristics of the first 492 patients with confirmed COVID-19, our study found that the number of male patients was higher (54%) than female patients (46%), consistent with several studies reporting from various parts of the world and indicating that men have higher morbidity and mortality associated with COVID-19 than women [15-17]. The male vs. female differences may be attributable to biological differences, including of the immune system, which impacts the body’s ability to fight an infection, including SARS-2-CoV-2. It has been reported that women are more resistant to infections than men, and this is potentially mediated by several factors, including different sex hormone levels and higher expression of coronavirus receptors (angiotensin-converting enzyme, ACE receptors) in men but also lifestyle aspects, such as higher levels of smoking and drinking among men compared with women [17]. Another vulnerability factor observed in our study was age, with the highest percentage of patients with confirmed COVID-19 being older than 60 years (20.4%), followed by patients aged 31 to 40 years (19.9%) and those aged 41 to 50 years (17%), a finding consistent with several other studies. There is evidence that the susceptibility to COVID-19 and the increased mortality risk in older people is linked to frailty [18]. Regarding clinical characteristics, we found that most symptomatic patients did not present with fever higher than 38 °C (54.8%) or dry cough (51.6%), and only 21% reported myalgia fatigue and 23.6% had abnormal findings on chest radiography. The latter finding was expected because COVID-19 may cause pneumonia that manifests clinically even in asymptomatic patients [19].

Among 492 patients, 21% were admitted in ICU attention, and 12% received mechanical ventilation. Approximately 42% of patients had comorbidities, including 20% with diabetes and 24% with hypertension. These results agree with a recent study from Spain in which they retrospectively described 49 consecutive patients admitted to the internal medicine hospital ward for COVID-19 infection and found a significant association between diabetes and the need for admission to the ICU [20]. One of the largest studies of this type, conducted in 138 hospitals in France, Belgium, and Switzerland and including over 4000 patients critically ill with COVID-19 admitted to an ICU, reported that patients who were older or had diabetes or obesity were at the highest risk of mortality [21]. In addition, our data showed patient outcomes at day 14 included persistent disease (33%), recovered (16%), discharged (8.4%) or death (1.5%). Our work also investigated variations in incubation time and its association with different variables. We found that the median incubation period reported in our population was 16 days (SD, 17 days), the upper 95% quartile was 27 days, and the lower quartile was 10 days. The incubation period was significantly associated only with age, with younger patients reporting a lower incubation period. Other investigated variables were not significantly associated with the incubation period. A systematic review and meta-analysis was recently published evaluating epidemiologic parameters to determine transmission and incubation dynamics [22]. For more than 23 studies combined, the mean incubation period of COVID-19 ranged from 4.8 to 9 days.

The present study determined whether patient profiles differed significantly for those who were symptomatic vs. asymptomatic. In at total of 429 patients, 79.2% reported having symptoms, and 20.7% reported being asymptomatic. Not every asymptomatic patient went to the hospital to be checked for the presence of SARS-CoV-2, which may explain the discrepancy between mild vs. severe patient outcomes. According to the WHO, most people infected with the SARS-CoV-2 virus will experience mild to moderate respiratory illness and recover without requiring special treatment; older people, and those with underlying medical problems, such as diabetes, chronic respiratory disease, cardiovascular disease, and cancer, are more likely to develop serious illness [23]. The age group and sex having the highest proportions of asymptomatic patients were 31-to-40-year-olds and males. Compared with symptomatic patients, asymptomatic patients were younger, nonsmokers, received an influenza vaccine, had no history of comorbidities, including diabetes or hypertension, and had lower BMI, ALT, and AST levels. Asymptomatic patients also had a higher NTL ratio and higher lymphocyte, CD4, CD8, natural killer cell, IgG, and IgM levels.

This study reported the most important epidemiologic and clinical factors associated with mortality in a cohort of patients with COVID-19. Our data showed that age and comorbidities were significantly associated with higher risk of death in this cohort, with both diabetes mellitus and hypertension associated with higher risk of death. Previous studies have shown that populations over 65 years old with comorbidities such as diabetes or hypertension have higher mortality rates in COVID-19 cases. The largest study of COVID-19 cases from China (72,314 cases) showed increased incidence of mortality among patients with diabetes and COVID-19 (2.3% without diabetes vs. 7.3% with the disease) [24]. Other studies from the United States, Italy, and China have reported that the diabetic population is at greater risk not only for disease complications but also for infection susceptibility [25].

Although an effective antiviral treatment for COVID-19 is not currently available, some repurposed medications have been proposed for use at KFSHRC. We observed different patient outcomes associated with each treatment regimen. Patients who did not receive an antiviral regimen had better prognoses, followed by patients receiving an interferon regimen. A significant and high HR was observed for patients receiving a combination therapy of azithromycin, hydroxychloroquine, and lopinavir/ritonavir, and a lower HR was observed for patients receiving a combination therapy of two or more drugs. Among patients who received only a single drug regimen, those receiving lopinavir/ritonavir or azithromycin had the worst prognosis. Our findings are consistent with the reported international data and clinical trials [26-29]. Patients receiving pneumonia treatment had poor prognosis overall, and patients receiving mechanical ventilation had also the second-highest rate. We concluded from our data on the treatment regimens that azithromycin, hydroxychloroquine, and lopinavir/ritonavir were not effective treatment for COVID 19, and this is consistent with previous studies and the solidarity trial [30, 31]

There are limitations in the current database and the main limitation is that this retrospective study was conducted during the early COVID-19 pandemic where various effective interventions were not assessed such as dexamethasone, IL6 inhibitors, Jak inhibitors and Remdesvir.

## Conclusion

This study reports significant differences between patients symptomatic and asymptomatic for COVID-19 in a cohort with a relatively large number at a Saudi tertiary care hospital, improving the understanding of the epidemiologic and clinical features associated with SARS-CoV-2 infection. This study also assesses treatment regimens used early in the COVID-19 pandemic to inform scientific and medical databases and future clinical use and research.

## Data Availability

The raw data supporting the conclusions of this article will be shared by the authors when requested.

## Contribution to the Field

This work provides epidemiologic and clinical characteristics data from patients in Saudi Arabia with confirmed SARS-COV-2 infection to inform treatment regimens and case management. Scarce studies from Saudi Arabia have highlighted case management and treatment regimens, especially for symptomatic vs. asymptomatic patients. The data provided here were obtained from a comprehensive database developed in-house for King Faisal Specialist Hospital and Research Centre, a single-center, tertiary and referral care hospital. We created the electronic medical records database to collect from each patient their demographic characteristics, virus exposure (travel history, and human and animal contact), vaccination history, comorbidities, signs and symptoms, laboratory and radiographic reports, cardiac workup, medications, treatment regimens, and outcome. The database continues to be used for all patients with confirmed SARS-COV-2 infection. Here, we describe our analyses of the demographic and clinical characteristic data obtained from the first 492 consecutive patients at the hospital diagnosed with SARS-CoV-2 infection. These findings provide insights on the epidemiologic and clinical characteristics and case management of patients in Saudi Arabia with confirmed SARS-COV-2 infection to inform future treatment regimens worldwide.

## Author Contributions

FA is the principal investigator of this study. RA and ED are co-principle investigators who helped in the study design and validation. BA, SA, MM, and AA are physicians and clinical scientists who reviewed medical charts and helped in clinical data interpretation. DO performed the statistical analysis. AA and SA built the database, trained the data collectors, and oversaw data collection and data quality. All authors helped in writing and reviewing the manuscript. In addition, all authors read and approved the final manuscript.

## Acknowledgements

We thank the KFSHRC higher administration for their full support during the pandemic. We also thank the data collection team who is managing the hospital database [Haya Alajlan, Ruba Aldossary, Samar Almogbel, Noura Alqahtani, Shatha Alshammari, Rahaf Al-Rashoud, Sanaa Al-Ali, Hadeel Al-Bedewi, Leen Al-Hogbani, Fai Al-Khathlan, Najla Al-Orayyidh, Noura Al-Zannan, Raneem Al-Zeer, Shatha Barajaa, and Ishraq Rajeh].

